# Simultaneous classification of bilateral hand gestures using bilateral microelectrode recordings in a tetraplegic patient

**DOI:** 10.1101/2020.06.02.20116913

**Authors:** Tessy M. Thomas, Robert W. Nickl, Margaret C. Thompson, Daniel N. Candrea, Matthew S. Fifer, David P. McMullen, Luke E. Osborn, Eric A. Pohlmeyer, Manuel Anaya, William S. Anderson, Brock A. Wester, Francesco V. Tenore, Gabriela L. Cantarero, Pablo A. Celnik, Nathan E. Crone

## Abstract

Most daily tasks require simultaneous control of both hands. Here we demonstrate simultaneous classification of gestures in both hands using multi-unit activity recorded from bilateral motor and somatosensory cortices of a tetraplegic participant. Attempted gestures were classified using hierarchical linear discriminant models trained separately for each hand. In an online experiment, gestures were continuously classified and used to control two robotic arms in a center-out movement task. Bimanual trials that required keeping one hand still resulted in the best performance (70.6%), followed by symmetric movement trials (50%) and asymmetric movement trials (22.7%). Our results indicate that gestures can be simultaneously decoded in both hands using two independently trained hand models concurrently, but online control using this approach becomes more difficult with increased complexity of bimanual gesture combinations. This study demonstrates the potential for restoring simultaneous control of both hands using a bilateral intracortical brain-machine interface.

## INTRODUCTION

Bimanual upper-limb control is crucial for completing many activities of daily living (ADLs). Simple tasks such as opening a bottle or squeezing toothpaste onto a toothbrush require simultaneous but independent control of both hands. Brain-machine interfaces (BMI) are a promising new technology to restore such ADLs in paralyzed individuals. While several BMI studies have demonstrated that paralyzed individuals can achieve successful unimanual reach and grasp control (Aflalo et al., 2015; Ajiboye et al., 2017; Bouton et al., 2016; Colachis et al., 2018; Collinger et al., 2013; Hochberg et al., 2012; Wang et al., 2013; Wodlinger et al., 2015; Yanagisawa et al., 2011, 2012), much less is known about the degree of bimanual control possible and how it can be affected by task complexity. Ifft et al. (2013) provided early evidence that bilateral arm movements can be decoded in real-time (online) from neural activity recorded through microelectrode arrays (MEAs) in bilateral sensorimotor cortices of non-human primates. More recent investigations have demonstrated that tetraplegic patients can also achieve simultaneous and independent 3D endpoint control of two arms using neural recordings from MEAs (Downey et al., 2019) and electrocorticography (ECoG) (Benabid et al., 2019).

However, simultaneous control of bilateral hand movements or gestures has not yet been demonstrated. Several studies have provided evidence that neural activity in one hemisphere, including single/multi-unit activity (SUA/MUA) and ECoG activity, contains adequate information for both discrete and continuous decoding of unimanual hand and finger movements (Acharya et al., 2010; Aggarwal et al., 2008; Bleichner et al., 2014; Bouton et al., 2016; Chestek et al., 2013; Colachis et al., 2018; Degenhart et al., 2018; Flint et al., 2017; Hamed et al., 2007; Hotson et al., 2016; Irwin et al., 2017; Jiang et al., 2017; Jorge et al., 2019; Liang & Bougrain, 2012; Nakanishi et al., 2014; Pan et al., 2018; Pistohl et al., 2012; Schwemmer et al., 2018; Vaskov et al., 2018). Wisneski et al. (2008) also demonstrated the ability to decode repeated opening and closing of either the contralateral or ipsilateral hand in real-time from ECoG signals recorded from one hemisphere in able-bodied subjects. Using microelectrode recordings, Downey et al. (2019) showed that right and left hand power grasps were difficult to differentiate from each other using MUA recorded from only one hemisphere in a tetraplegic subject. Instead, they found a high correlation between the neural tuning to these grasps within the motor cortex of one hemisphere. Recording from both hemispheres could potentially improve independent and simultaneous control of bilateral hand movements for patients with spinal cord injuries (SCI). Notwithstanding the added risks from bilateral implants, they have potential advantages over unilateral implants for people with intact sensorimotor cortex in both hemispheres, including SCI patients. One group showed preliminary evidence that simultaneous opening and closing of both hands can be decoded from bilateral sensorimotor activity recorded through magnetoencephalography in able-bodied subjects (Belkacem et al., 2018). Using a more invasive recording method, Benabid et al. (2019) used bilateral ECoG recordings to provide a tetraplegic patient with control of translation and wrist rotation of an exoskeleton’s two upper-limbs. However, the degree of complex and dexterous bilateral movements achievable with bilateral MEA recordings in a human subject has not yet been investigated.

As part of an ongoing early feasibility study of bilateral intracortical BMIs for controlling bilateral upper-limb movements, we implanted MEAs in the sensorimotor cortex of both hemispheres in an SCI subject with incomplete tetraplegia. Here, we present the results of simultaneous online classification of bilateral hand gestures. Bimanual gesture combinations were predicted using two hierarchical linear classifiers, each trained for a specific hand using inputs from the contralateral motor and somatosensory cortices. Furthermore, by mapping a set of hand gestures to four directions on a 2D plane, the participant was able to use the gesture classifiers to simultaneously control two JHU/APL Modular Prosthetic limbs (MPLs; Johannes et al., 2011, 2020) in center-out movement tasks. Online performance was explored as a function of the complexity of bilateral gesture combinations. This study represents the first demonstration of simultaneous gesture decoding in both hands using bilateral microelectrode recordings.

## RESULTS

### Offline Unilateral Gesture Classification

A 48-year-old male participant with incomplete tetraplegia (C5/C6, ASIA B) was implanted with two pairs of MEAs within the left hemisphere and one pair of MEAs within the right hemisphere. Each pair consisted of a 96-channel array in the motor cortex and a 32-channel array in the somatosensory cortex (Figure 1). Starting from day 189 post-implant, the participant completed 14 gesture classification sessions over three months. Each session began with a training task, in which the participant was visually cued to attempt unimanual gestures in a randomized order. A hierarchical classification model was trained for each hand by using linear discriminant analysis (LDA) on MUA recorded from only the contralateral hemisphere. This hierarchical framework consisted of a linear model trained to first choose between rest and movement (first step), and a second linear model trained to predict a specific gesture (second step).

**Figure 1.**
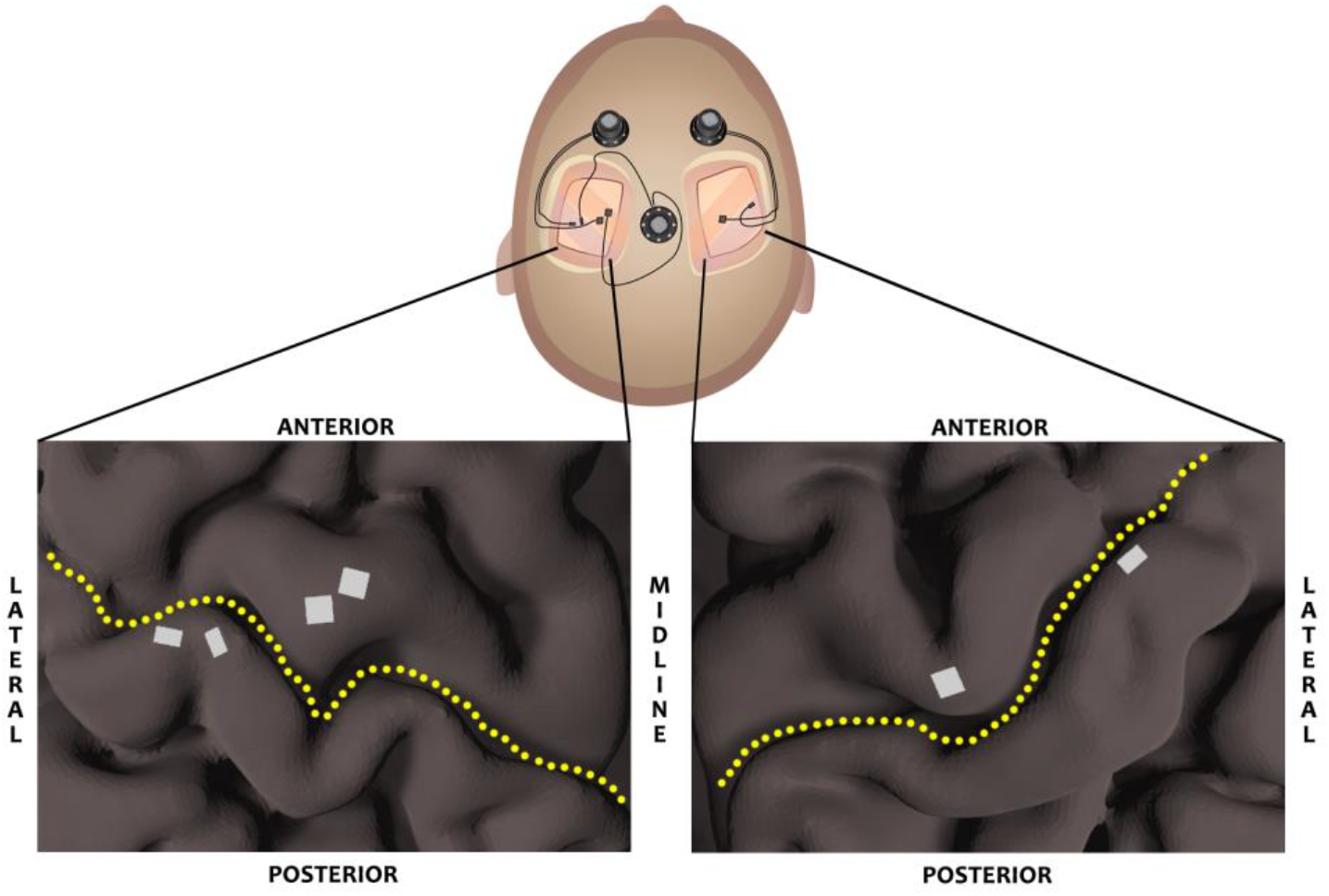
MEA implant locations. 6 MEAs were implanted in pairs, with a 96-channel array in the motor cortex and a 32-channel array in the somatosensory cortex. Each pair was connected to a pedestal on top of the skull (top image), through which neural signals were recorded. Two pairs of MEAs (shown in white) were implanted in the left hemisphere (left image). One pair of MEAs was implanted in the right hemisphere (right image). Yellow dotted-line outlines the central sulcus.

**Table 1.** Offline gesture classification accuracies. Average classification accuracies for rest and eight attempted unilateral gestures for each hand across 14 sessions.

|  | Rest | Power Grasp | Open Hand | 2-Finger Pinch | 5-Finger Flat Pinch | Wrist Flex | Wrist Extend | Index Flex | Index Extend |
| --- | --- | --- | --- | --- | --- | --- | --- | --- | --- |
| Right Hand | 93.3% | 73.0% | 63.6% | 81.3% | 73.0% | 76.2% | 75.5% | 75.0% | 65.0% |
| Left Hand | 89.4% | 33.0% | 53.7% | 70.3% | 57.7% | 67.6% | 57.7% | 45.0% | 35.0% |

Table 1 shows the average offline classification accuracy of each gesture which was used for at least one online task (Figure S1). The first step of the hierarchical model classified between rest and movement with an accuracy of 93.3% and 89.4% for the right and left hand, respectively. Right hand gestures, decoded in the second step, were more accurately classified than left hand gestures (Wilcoxon rank-sum test, p=1.78×10^-6^). Among the gestures, 2-finger pinch was most accurately classified (81.3% on right hand, 70.3% on left hand), followed by classification of wrist flexion and wrist extension. As a result, these three gestures were frequently used as movement classes for a majority of experimental sessions. Early sessions included power grasp as one of the gestures, but it was poorly classified on the left hand and was replaced with other gestures in later sessions. The alternative gestures included open hand, 5-finger flat pinch, index flex, and index extend, of which the first two were most consistently discriminable on both hands. Different gestures were substituted in order to use the most discriminable gesture set across both hands for online control during each session.

### Online Bilateral Gesture Classification

In order to assess online classification of gestures in both hands simultaneously, the participant used the gesture classifiers to drive two MPLs and perform a bimanual center-out movement task. Taking a similar approach to previous studies (Degenhart et al., 2018; Wang et al., 2013), a specific gesture was mapped to each of the four cardinal directions on a 2D vertical plane, and the predicted gesture from each hand’s model was used to guide the corresponding MPL at a constant velocity towards a target on the vertical or horizontal axis. The same gestures were used for both hands, and the directional mapping was mirrored across both hands (Figure S2). This task provided an experimentally controlled environment to test the effectiveness of the gesture classifiers for online control while also keeping the subject engaged in achieving a goal. The bimanual task required the participant to use a hierarchical classification model for each hand simultaneously in order to independently control both MPLs. Bimanual trials consisted of three types of bimanual gesture combinations: stabilizing trials (also referred to as “Gesture+Stabilize”: one model must predict a gesture while the other model predicts “rest” simultaneously), symmetric gesture trials (both models must simultaneously predict the same gesture), and asymmetric gesture trials (both models must simultaneously predict different gestures). A unimanual center-out movement task, in which only one model and one MPL were active during each trial, was also performed to obtain a benchmark for comparing two-hand gesture classification. Unimanual trials consisted of two types: isolated unimanual trials (used only one hand’s model during all trials in a block), and alternating unimanual trials (switched between the right and left hand models for each trial in a block).

The participant completed 32 blocks of the unimanual task and 16 blocks of the bimanual task. The average success rate for reaching the target (target-reach) was 76% for unimanual blocks and 63.8% for bimanual blocks (Figure 2A). The distribution of target-reach success rates for the unimanual task was significantly higher than the distribution for the bimanual task (Wilcoxon rank-sum test, p=0.016). But with a median trial-completion time of 3.19 s (unimanual) and 3.07 s (bimanual), there was no significant difference (Wilcoxon rank-sum test, p=0.49) in the time it took to successfully reach the target between the two tasks (Figure 2B).

**Figure 2.**
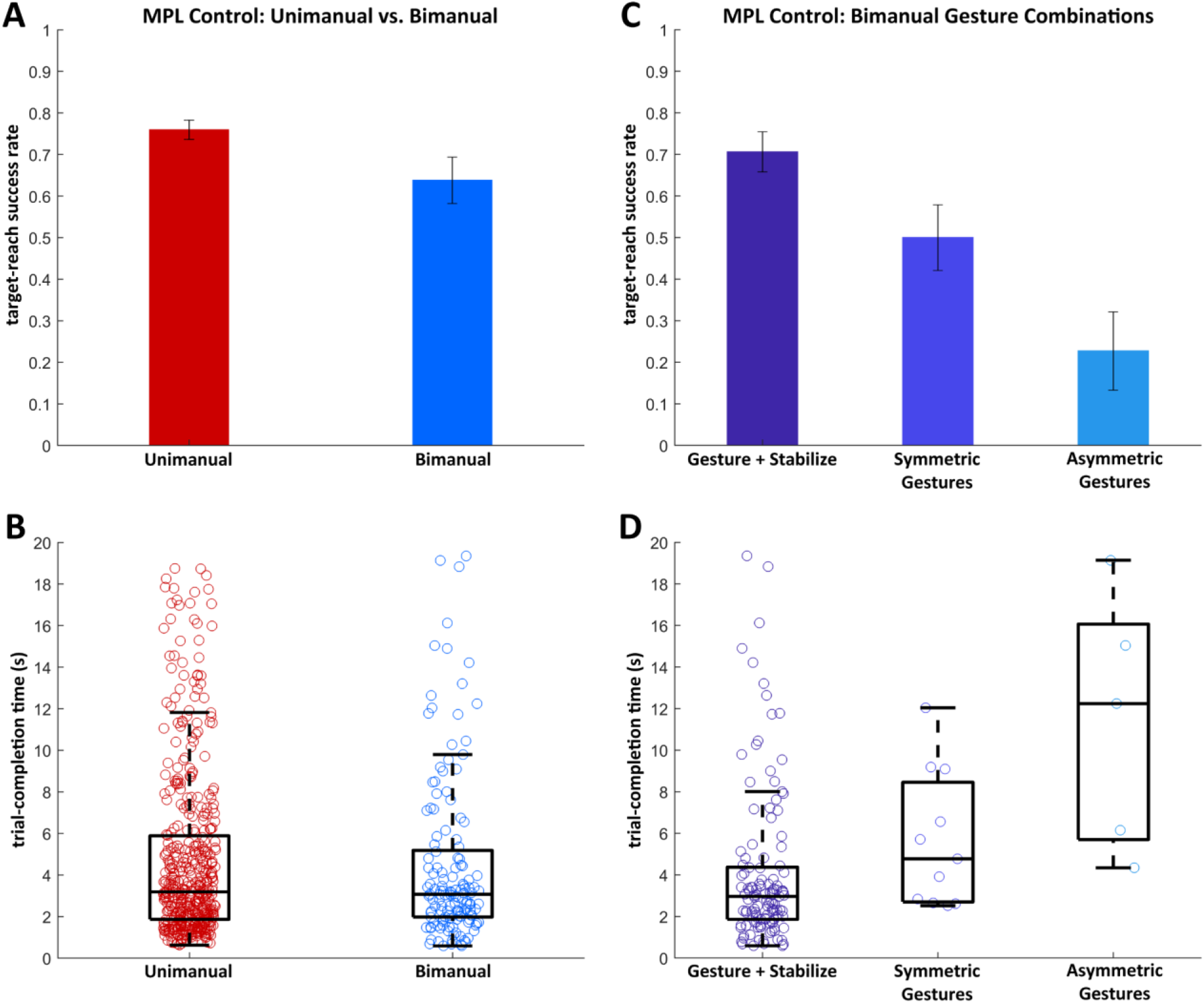
Online center-out task performance using gesture classification. (A) Average success rate and (B) trial-completion time for reaching targets (target-reach) using predicted gestures are shown for unimanual trials and bimanual trials, as well as for three different bimanual combinations (C, D). Error bars signify standard error. The time it took to successfully reach the target during individual trials (trial-completion times) is plotted as vertical scatterplots for each task type (B, D). Boxplots display a summary of the distribution of the underlying scatterplot. The edges and middle line of the box represent 25^th^ percentile, median, and 75^th^ percentile from bottom to top. Circles that fall outside of the boxplot whiskers are outliers.

Trials from the bimanual tasks were further divided into three groups according to the complexity of bimanual gesture prediction. These groups included stabilizing trials, symmetric gesture trials, and asymmetric gesture trials. Each of these bimanual trial groups required simultaneous classification of gestures on both hands with varying degrees of independence and difficulty, with stabilizing trials as the least complex and asymmetric trials as the most complex gesture combination. The participant achieved a higher target-reach success rate for the stabilizing trials (70.6%) than for the symmetric and asymmetric gesture trials (Figure 2C). Symmetric gesture trials were completed with an average target-reach success rate of 50%. Performance was poorest for asymmetric gesture trials, in which the participant successfully reached both targets during only 22.7% of the trials. Stabilizing trials also had the shortest median trial-completion time, followed by symmetric and asymmetric gesture trials (Figure 2D).

### Right vs. Left Hand Performance

We tested for significant differences in performance between the right hand and left hand models (“significant effects of handedness”) in terms of target-reach success rates and trial-completion times during three trial types: isolated unimanual trials, alternating unimanual trials, and bimanual stabilizing trials. Bimanual symmetric and asymmetric gesture trials were excluded from this comparison because both hands were required to achieve their goals simultaneously, resulting in the same target-reach success rates and trial-completion times for both right hand and left hand models. The average target-reach success rate achieved by the right hand model was 3.8%, 11.1%, and 2.6% higher than that achieved by the left hand model for the isolated unimanual, alternating unimanual, and bimanual stabilizing trials, respectively (Figure 3A). Median trial-completion times, ranging between 2.81 s and 3.45 s, were very similar for both the right and left hand models across all three trial types (Figure 3B). Even though the right hand model had slightly higher target-reach success rates than the left hand model across all three trial types, there were no significant effects of handedness or trial type on either target-reach success rate (p=0.20, p=0.06, two-way ANOVA) or trial-completion time (p=0.53, p=0.093, two-way ANOVA).

**Figure 3.**
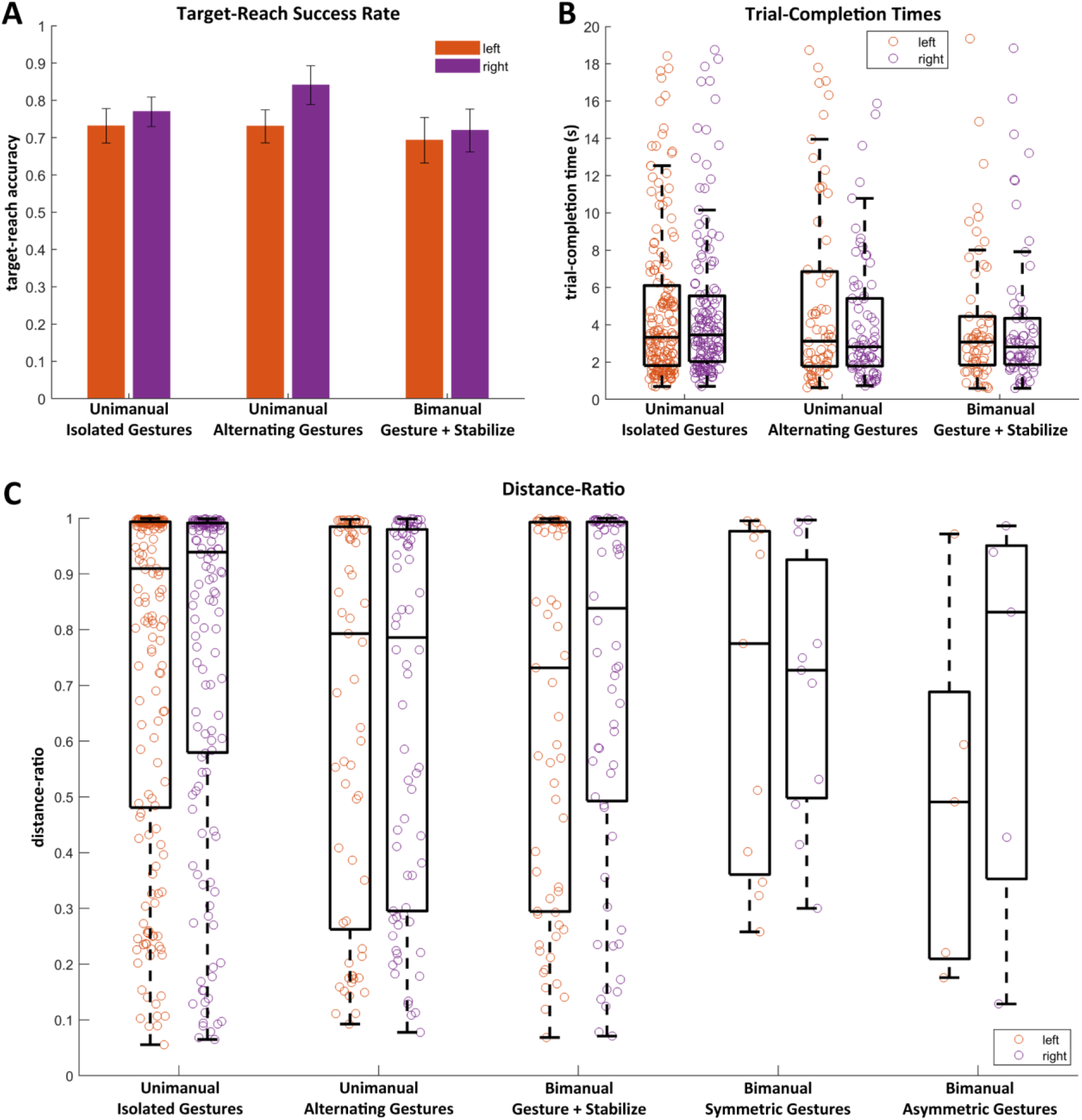
Online performance of right vs. left hand. (A) Average target-reach success rates and (B) distributions of trial-completion times for the right and left hand are shown for two unimanual and one bimanual task types. (C) Distance-ratio (the straightest and shortest distance to the target divided by the distance traveled by the MPL to the target) for successful trials is plotted as vertical scatterplots and overlaid with boxplots for right and left hand for all five task types.

We used distance-ratio, defined as the straightest and shortest distance to the target relative to the actual distance traveled by the MPL (from worst to ideal ranging from 0 to 1), as an additional performance metric to compare right and left hand models across all five different trial types. The median distance-ratio for isolated unimanual trials was the closest to the ideal ratio. However, there was much variance among the distance traveled in individual trials for all trial types, as shown by the wide distribution of distance-ratio values (Figure 3C). Handedness did not have any significant effect on distance-ratio (p=0.28), but trial type did have a significant effect (p=0.016, two-way ANOVA).

### Motor vs. Sensory Contributions to Decoding

The model weights used to discriminate between attempted gestures during the second step of the hierarchical classifiers were analyzed to determine the decoding contributions of motor and sensory electrodes (Figure 4). In the left hemisphere, 47% out of 192 motor electrodes and 100% out of 64 sensory electrodes were used by the right hand model during at least two sessions. One electrode on the lateral sensory MEA and one electrode on the medial motor MEA were the most informative. The motor electrodes that contributed to the model were primarily located on the medial motor MEA, with a small cluster of electrodes providing relatively high decoding power (> 0.6). The minimal contribution (< 0.16) from the lateral motor MEA was likely due to an overall low yield of SUA and MUA from that array. Since the time of implantation, we have observed almost no modulation in this array during a wide variety of attempted and executed proximal and distal upper-limb movements. The lateral sensory array contributed more to decoding than the medial sensory array. Based on pre-implant functional mapping, this array is likely situated near somatosensory thumb and index representations, which may have helped discriminate between gestures such as pinch and open hand. In the right hemisphere, 67% out of 96 motor electrodes and 91% out of 32 sensory electrodes were used by the left hand model during at least two sessions. The most informative electrode was located on the motor MEA and was part of a cluster of electrodes along the lateral edge of the array with relatively strong contributions (> 0.73) to decoding. The sensory MEA also showed high decoding power, though not as uniformly as its homologous array in the opposite hemisphere. Across motor and sensory arrays in both hemispheres, the electrodes with higher decoding power were more consistently included in the model across multiple sessions.

**Figure 4.**
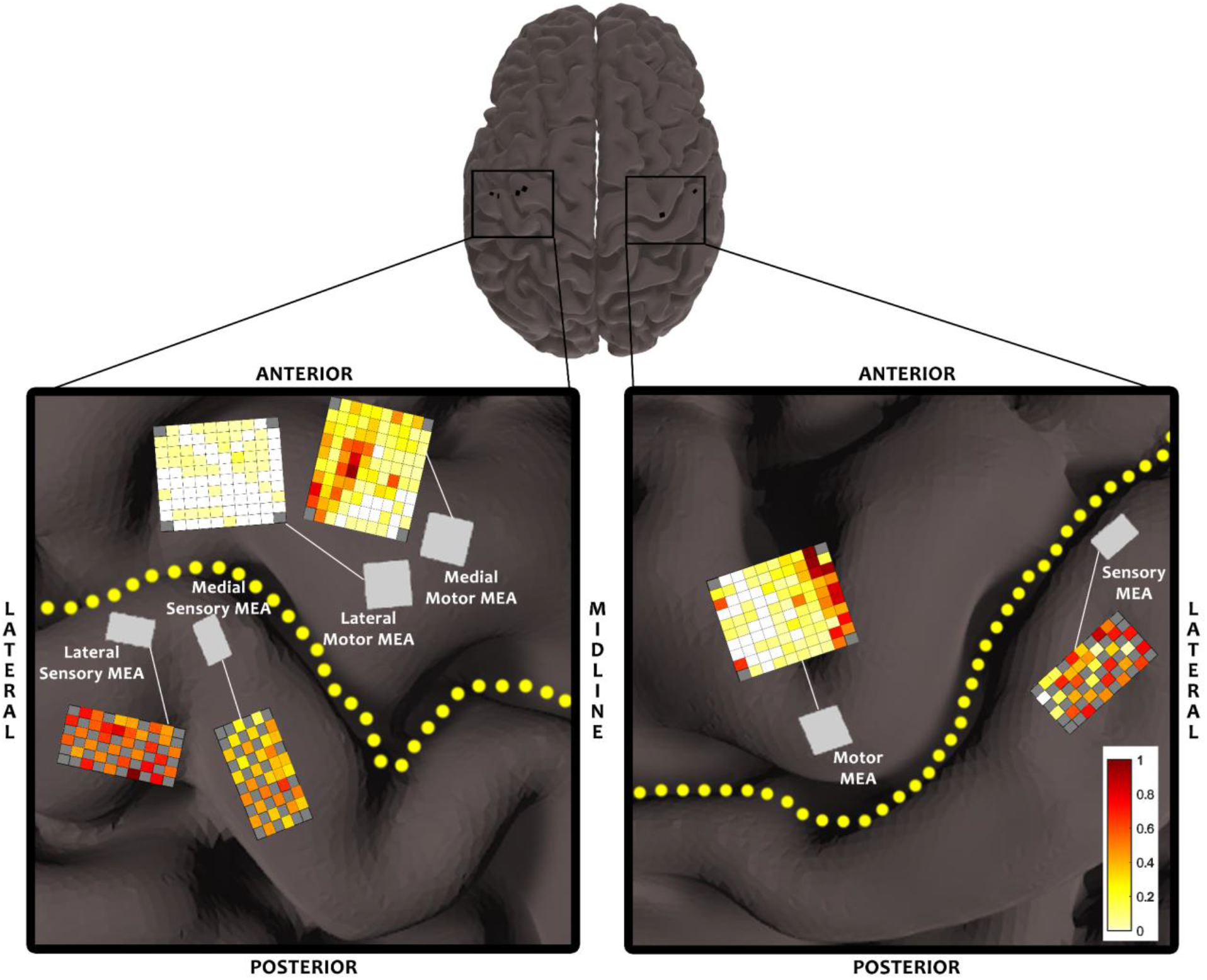
Decoding contributions of motor and sensory MEAs. Normalized weights of each electrode in the gesture classification models are displayed by a color ranging from light yellow (least contributing) to dark red (most contributing). White electrodes were not included in the classification model (see MATERIALS AND METHODS). Decoding weights of electrodes in each hemisphere were normalized to the highest weight observed across all motor and sensory arrays in that hemisphere. Arrays on the left hemisphere (left image) were used to classify right hand gestures, and arrays on the right hemisphere (right image) were used to classify left hand gestures. Yellow-dotted line outlines the central sulcus.

## DISCUSSION

Most daily tasks require simultaneous use of both hands for grasping and interacting with one or more objects. In this study, we demonstrated, for the first time, online classification of bilateral hand gestures using multi-unit activity recorded from bilateral motor and somatosensory cortices in an SCI patient with incomplete tetraplegia. Not only were movements for both hands classified at the same time, but they were also classified independently of each other. Simultaneous and independent classification may have been possible because an independent decoding model was constructed for each hand by restricting the model inputs to MUA in only the contralateral hemisphere. In light of the observation of Downey et al. (2019) that MUA from the motor cortex in one hemisphere contains correlated neural representations of right and left hand grasps, it is possible that using microelectrode recordings from both hemispheres also reduces interference from the ipsilateral hand within each hemisphere.

While we showed that it was possible to classify bimanual gesture combinations, we found that online performance tended to decrease as bimanual combinations became more complex. Our decoding model performed well when the participant attempted movements on one hand while simultaneously keeping the other hand at rest, but had difficulty in decoding symmetric and asymmetric gestures, with the poorest performance observed for simultaneous asymmetric gestures. These differences in bimanual decoding performance could have been due to different demands on attention. For example, more complex gesture combinations may have required more divided attention between the two hands when attempting to control both MPLs, leading to a decrease in online performance. When executing a movement with one hand while continuously maintaining the other hand at rest, attention can focus primarily on the moving hand while the resting hand is passively monitored. Symmetric movements may have required more attention than stabilizing movements, but may have still been feasible due to similar movement kinematics across both hands. In contrast, asymmetric movements involved simultaneous attention to independent goals for both hands. Given that such asymmetric tasks can be difficult for most people, it may not be surprising that online performance was worse during asymmetric gesture trials than during symmetric gesture trials. Another potential reason why our decoding models generalized better for stabilizing trials is that the classification models for each hand were trained solely on unimanual gestures, for which attention and kinematics were very similar to the stabilizing trials. In contrast, symmetric and asymmetric bimanual gesture combinations may have had neural representations that were very different from unimanual gestures, possibly because of neural activity encoding both contralateral and ipsilateral hand movements in the same hemisphere. Several studies have provided evidence that contralateral, ipsilateral, and bilateral reaching and finger movements can evoke neural activations in the same hemisphere (Aizawa et al., 1990; Ames & Churchland, 2019; Cisek et al., 2003; Cramer et al., 1999; Diedrichsen et al., 2013; Donchin et al., 1998, 2002; Heming et al., 2019; Kim et al., 1993; Steinberg et al., 2002). If models for each hand are trained on both unimanual and bimanual gestures, they could potentially generalize to a wider range of bimanual gesture combinations. However, to train on all possible bimanual combinations would have greatly expanded the time required for training.

When we compared the performance of independent right and left hand models, we observed a significant difference in offline classification accuracies, namely higher accuracy for right hand gestures. Right hand models may have achieved a higher accuracy due to having more inputs than the left hand models. With twice as many electrodes in the left hemisphere as the right hemisphere, the right hand model may have been more capable of capturing separable neural representations for the different gestures. In particular, the right hand model received twice as many somatosensory inputs as the left hand model, and the additional decoding power from these extra inputs may have resulted in a higher classification accuracy for the right hand. Interestingly we did not observe this same disparity between right hand and left hand models when comparing online performances. The lack of any significant differences between the two hands may have resulted from the specific nature of the online task. Offline assessment required the model to classify gestures from only a single attempt. In contrast, the online assessment involved using the model’s predictions to drive the MPLs toward targets that were far enough from the center of the movement space to require multiple attempts of a gesture. Repeated gesture attempts may have resulted in a more consistent increase in firing rates, which could have yielded higher and more similar classification accuracies for the two hands during the online task. Further studies will be needed to test whether this similarity in online performance will extend to scenarios in which classification models are used to execute hand and wrist movements with the MPLs.

Both motor and somatosensory cortices in the contralateral hemisphere contributed to decoding gestures in each hand. In each hemisphere, the proportion of informative motor electrodes was smaller than the proportion of informative sensory electrodes. It is possible that cortical activation in the “hand knob” area was significantly reduced as a result of the participant’s SCI, resulting in sparse areas of activation difficult to capture within a single MEA. The motor electrodes with the highest decoding power also tended to be clustered to one side of the MEA, which suggests that the arrays captured one edge of the neural representations for relevant hand movements. The lateral motor MEA in the left hemisphere may have been in a more ideal location to capture these neural representations, but the low yield of SUA and MUA from that array may have led to its minimal contribution to decoding. Somatosensory cortical areas associated with individual finger sensations also showed high decoding power, which may have aided in differentiating gestures with opposing finger positions such as pinch, power, and open hand. Since the participant retained partial sensation in both hands, some of the neural activity in somatosensory cortex may have originated as sensory feedback from slight finger movements during attempted gestures. This activity could have also resulted from movement-related information received from the motor cortex, such as an efference copy (Crapse & Sommer, 2008; Favorov et al., 2015; Lebedev et al., 1994; Soso & Fetz, 1980). While sensory feedback may not be present in other individuals with more complete SCI, informative somatosensory activations could result from cortical inputs from motor areas. However, due to the higher prevalence of incomplete SCI than complete SCI among the affected population, these results could generalize to a broader population.

An important step to restoring complete bimanual control through a BMI is to provide the ability to control both hands at the same time. Here we show that it is possible to simultaneously and independently decode hand gestures in both hands from bilateral MEA recordings. These results suggest that a BMI user can utilize two classification models together to independently control two end-effectors. This approach could potentially be used to restore bimanual grasp control to SCI patients through functional electrical stimulation of their muscles or control of a bilateral exoskeleton, or to bilateral upper-limb amputees through control of two neuroprostheses. Further investigations will be needed to gain a better understanding of the neural representations of increasingly complex bimanual hand movements and their interactions across the two hemispheres. The ability to achieve simultaneous control of complex and dexterous movements in both hands could expand the activities of daily living possible for these individuals and increase their independence.

## MATERIALS AND METHODS

### Research Participant

In a study conducted under an Investigational Device Exemption (170010) by the Food and Drug Administration and approved by the Johns Hopkins Institutional Review Board and NIWC Pacific IRB, an incomplete tetraplegic (C5/C6, ASIA B) participant was recruited through a registered clinical trial (NCT03161067) and implanted with six MEAs 30 years post-injury. The participant had some retained motor control of the upper arm and wrist extension on both sides but had little to no movement in the rest of his hands and fingers. Pre-operative examination revealed intact light touch sensation in hands and fingers, but found some deficits to pinprick sensation in certain areas.

### Implantation Procedure

Two 96-channel MEAs (Pt-tipped Utah arrays – Blackrock Microsystems, Salt Lake City, Utah) were placed in the left primary motor cortex (M1), and one 96-channel MEA was placed in the right M1 (Figure 1). Two 32-channel MEAs (SIROF-tipped) were placed in the left primary somatosensory cortex (S1), and one 32-channel MEA was placed in the right S1 (Figure 1). These arrays were capable of both stimulating and recording from neural units. Pre-operative 7T structural and functional MRI (fMRI) were used to target the placement of these arrays within the pre-central and post-central gyri in both hemispheres. fMRI activation maps of physical and attempted movements of the hand, wrist, elbow, and shoulder highlighted the most active areas of the hand knob. Similarly, fMRI activation maps from mechanical stimulation of individual fingers highlighted finger representations in the post-central gyrus. Additionally, high-density ECoG functional mapping was performed intra-operatively to localize the representations of individual fingers within the cortical area informed by fMRI. High-gamma (70-110 Hz) power modulations were mapped in real-time from a 3×21 high-density ECoG grid (1mm contacts, 3mm spacing) placed on the post-central gyrus of one hemisphere during vibrotactile stimulation of individual fingers of the contralateral hand. The resulting finger activation maps helped to target the sensory MEAs in cortical areas that straddled the border between 2-3 finger representations.

### Experiment Description

During each session, the participant first completed a training task that involved attempting different gestures in a trial-by-trial manner. The gestures were randomly cued by showing the participant a picture of the gesture for 2 s followed by a 2-3 s pause. The gestures used across all sessions included power grasp, open hand, 2-finger pinch, 5-finger precision pinch, wrist flexion, wrist extension, index flexion, index extension, and rest (Figure S1). The participant was instructed to attempt the cued gesture to the fullest extent possible and hold the gesture until the picture disappeared from the screen. The participant’s upper arms and forearms were positioned in a neutral position, and the hands were held in a neutral position with gravity eliminated in-between trials (palms facing each other). The participant was able to physically execute certain gestures to some extent, such as wrist extension and wrist flexion, but could only produce minimal finger twitches when attempting other hand movements. After the training task was completed, classification models were trained and validated to report offline classification accuracies.

The participant then used the classifiers online to complete unimanual and bimanual center-out tasks. The set of gestures that yielded the best discriminability offline were mapped to four directions along the vertical and horizontal axes (Figure S2). The participant’s predicted gestures were used to guide each MPL at a constant velocity towards a target at the end of one axis. Each MPL had a separate set of four targets, and each MPL began at a “home” position located at the center of its respective 2D vertical plane. Once a target was displayed for one or both MPLs, the participant was instructed to move the MPL(s) to the respective target(s) in as straight a path as possible. The maximum time allowed for each trial was 20 s. Once the participant successfully reached the targets or the trial time exceeded this time limit, the MPLs were automatically reset to the home position.

### Neural Recording and Pre-processing

Neural activity was recorded from the MEAs using three 128-channel Neuroport Neural Signal Processors (Blackrock Microsystems). At the beginning of each session, the voltage threshold for each electrode was set at −3.5 times the root-mean-square voltage (RMS) of resting-state activity. Spiking activity was captured at 30 kHz, and the multi-unit firing rate for each electrode was calculated by counting the number of spikes that crossed the threshold within each 30 ms bin. The square-root of the firing rate at each time point was normalized to the mean and standard deviation (z-score calculation) of the square-rooted firing rates within the previous 60 s.

### Offline Classifier Training and Validation

Normalized firing rates recorded from both motor and sensory electrodes within each hemisphere during the training task were segmented and used to build a classification model for the contralateral hand. The average normalized firing rate of each electrode within a 1 s window relative to cue-onset of each trial was calculated. The start of the window was shifted forward by 250 ms or 500 ms from cue-onset to account for the participant’s reaction time and to capture neural activity corresponding to movement attempt. The amount of shift was determined from offline visualization of the average neural activity aligned to cue-onset for each attempted gesture. Computing the average firing rate for each electrode resulted in a set of 384 neural features (combined number of electrodes across all six arrays) for each trial. An ANOVA test was used to identify the electrodes that showed a significant difference in average firing rate between at least two gesture classes (p<0.05, FDR corrected). The neural features corresponding to the selected electrodes were used to train a hierarchical classification model using linear discriminant analysis. The first step of the model was trained to classify between rest and movement, while the second step was trained to classify between the different gestures. The second step of the classifier would only be used if the first step predicted movement. A separate classifier was built for each hand using neural features from only the contralateral hemisphere. Each model was trained and validated using 10-fold cross-validation.

### Online MPL Control

Online control was tested using unimanual and bimanual center-out tasks. The participant used only the classifier corresponding to the active MPL during each trial of a unimanual task, and used two classifiers to control both MPLs simultaneously during each trial of a bimanual task. The online decoder received binned spike counts every 30 ms, which were pre-processed in the same way as the training data. Normalized firing rates from the electrodes were stored in a 1 s buffer and updated every 30 ms. The buffered firing rates were averaged and used by the hierarchical classification model to make a rest or gesture prediction. A prediction was made every 30 ms. For each block of a unimanual or bimanual testing task, online control was assessed by computing the target-reach success rate, trial-completion times, and distance-ratios. Target-reach success rate was determined as the number of trials in which the participant successfully navigated the MPLs to the cued targets divided by the total number of trials. A successful unimanual trial required the MPL to reach its target within 20 s and stay on target for 100 ms. A successful bimanual trial required both MPLs to reach their targets within 20 s and simultaneously be on their targets for 100 ms. Trial-completion time was determined for each successful trial as the time it took to reach the target. Distance-ratio was also calculated for each successful trial as the straightest and shortest distance to the target divided by the actual distance traveled to the target. All three performance metrics were compared using a two-way ANOVA test to test for significant differences in the performance of right and left hand models within and across the different task types.

### Characterizing Cortical Contributions to Decoding

To analyze the relative decoding contributions of motor and sensory electrodes from each hemisphere to decoding gestures on the contralateral hand, the weights assigned to each electrode were extracted from the classification model. The weight vectors assigned to each electrode by a given classification model were summed together and normalized. All electrode weights from each hemisphere were normalized to the highest weight across all motor and sensory electrodes in that hemisphere. These normalized weights were displayed as a heat map ranging from 0 to 1 on the arrays.

## Data Availability

Data is not publicly available at this time.

## ACKNOWLEDGMENTS

The authors would like to thank our study participant for volunteering his time and energy to this study. This participant also contributed to two other studies (preprint submissions MEDRXIV/2020/117374 and MEDRXIV/2020/117036). The authors would also like to thank Rob Franklin, Stephen Hou, and the staff at Blackrock Microsystems for technical support during implantation and neural recording; Wouter Schellekens and Nick Ramsey for fMRI processing and analysis; Chad Gordon, Teresa Wojtasiewicz, Adam Schiavi, and the surgical team at Johns Hopkins Hospital for performing the implantation; Christopher Coogan for MRI reconstruction and software support of experimental paradigm implementation; Bryanna Yeh, Christian Cooke, and Zachary Koterba for development and support of the data-recording software infrastructure; and Christopher Dohopolski, Jared Wormley, John Roycroft, and Matthew Johannes for MPL infrastructure design and support. This research was developed with funding from the Defense Advanced Research Projects Agency’s (DARPA, Arlington) Revolutionizing Prosthetics program (contract number N66001-10-C-4056). The views, opinions, and/or findings expressed are those of the author(s) and should not be interpreted as representing the official views or policies of the Department of Defense or the U.S. Government. Development of experimental setup and support for regulatory submissions associated with this study were provided by a grant from the Alfred E. Mann Foundation. Study software infrastructure and study preparation were developed with internal funding from Johns Hopkins University Applied Physics Laboratory and Johns Hopkins University. Personnel was also supported by NIH (R01NS088606).

## SUPPLEMENTAL FIGURES

**Figure S1.**
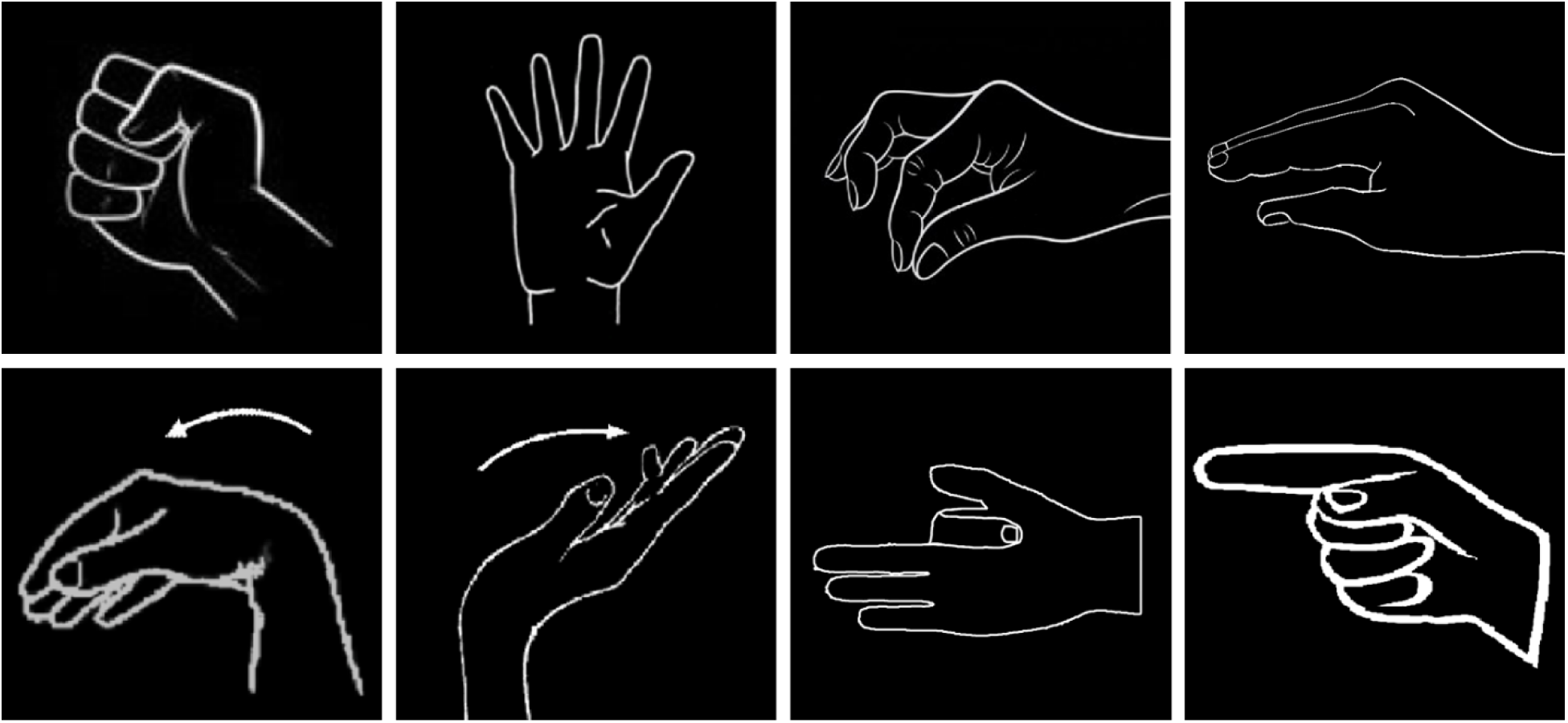
The different gestures used for training and testing hierarchical classification models across 14 sessions. Top row: power grasp, open hand, 2-finger pinch, 5-finger flat pinch. Bottom row: wrist flex, wrist extend, index flex, index extend. Right hand gestures are shown here. Left hand gestures (mirror versions of these images) were also shown during the training phase.

**Figure S2.**
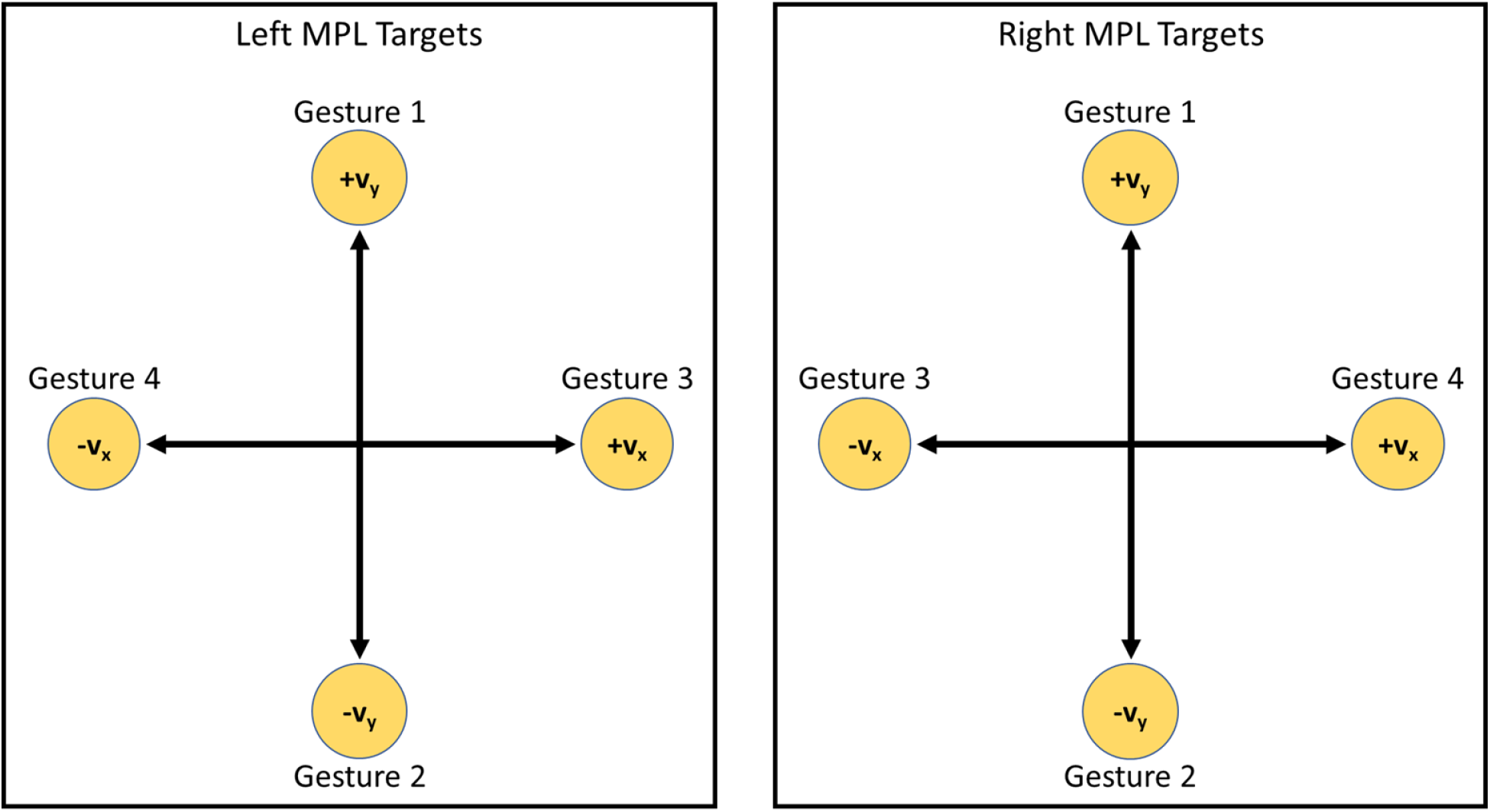
Four gestures were mapped to four cardinal directions in the vertical 2D plane. The same gestures were used to control the left and right MPLs, and the gesture-direction mapping was mirrored across the two MPLs.

